# AI Perspectives on the Present and Future of Antidepressant Pharmaceutical Treatment Based on Anti-inflammatory Strategies: A Scoping Review of Randomised Controlled Clinical Trials

**DOI:** 10.1101/2024.12.31.24319839

**Authors:** Yan Bo

## Abstract

**Background:** Depression remains an unresolved issue on a global scale. Recently, a novel concept of ‘anti-inflammatory-based pharmacotherapy’ has been developed. Despite the role of inflammation in depression having been discussed in many reviews at various levels, the prevalence of this new concept in randomised controlled clinical studies and its implications remain elusive. The aim of this scoping review was precisely to explore in depth the current status of inflammation in randomised controlled clinical trial studies of depression.

**Methods:** PubMed was systematically searched from inception to December 11 2024. Studies that researches on the treatment of depression based on anti-inflammatory strategies were included. Study characteristics and outcomes were extracted and organized thematically.

**Findings:** 11 reports of randomised controlled clinical trials were included, which accumulated 907 depressed patients. All studies found that there is a connection between the effects of anti-inflammatory drugs in treating depression and a large decrease in the levels of inflammatory markers in the blood of depressed patients compared to before treatment. Three inflammatory markers, CRP, IL-6 and TNF-alpha, were the most frequently mentioned. The current strategy of anti-inflammatory drug administration did not differ fundamentally from the previous strategy of traditional antidepressant drugs combined with psychotherapy.

**Interpretation:** At present, the use of anti-inflammatory strategies for the pharmacological treatment of depression has limited research value and poor feasibility. The future direction of the new concept of anti-inflammatory strategies for the treatment of depression, proposed in the context of the association between inflammation and depression, is that psychiatrists, researchers, and psychotherapists should shift their future focus from pharmacological treatments based on anti-inflammatory strategies to non-pharmacological treatments of anti-inflammatory strategies, such as positive thinking, exercise, and so on. The popularity of purely clinical randomised controlled studies in the depression population is extremely low when considering the financial investment in research and the benefits of translating the results. In the future, public interest studies, low research costs, and research protocols with mass generalisability will be more likely to stimulate the depression community’s interest in participating in research. The potential value and feasibility of future research lies in the application of an integrated AI platform to assist pharmacological treatment of depression based on anti-inflammatory strategies.

Registration DOI: https://doi.org/10.17605/OSF.IO/A64GC.

## 1 Research in context

### 1.1 Evidence before this study

Depression remains an unresolved issue on a global scale. Recently, a novel concept of ‘anti-inflammatory-based pharmacotherapy’ has been developed. Despite the role of inflammation in depression having been discussed in many reviews at various levels, the prevalence of this new concept in randomised controlled clinical studies and its implications remain elusive. My previous preliminary search of the PubMed database revealed no extant systematic reviews publishing insights on how to view anti-inflammatory strategies based on the treatment of patients with depression-associated inflammation.

### 1.2 Added value of this study

To the best of our knowledge, this is the first scoping review that investigates treatment of patients with depression-related inflammation based on anti-inflammatory strategies. It uncovers the connection between anti-inflammatory drugs, reduced inflammatory markers, and treatment effects. It also points out the limitations of current anti-inflammatory strategies in treating depression, suggesting a shift to non-pharmacological approaches. Additionally, it proposes the potential of AI in improving treatment, providing new directions for future research.

### 1.3 Implications of all the available evidence

Current evidence shows anti-inflammatory strategies for depression treatment have limitations. Future research should focus on non-pharmacological approaches, AI-assisted treatment, and clarifying inflammation-depression links for better patient care.

## 2 Introduction

In recent years, the role of inflammation in depression and the effects of anti-inflammatory drugs on depression have emerged as significant areas of mental health research. A growing body of evidence strongly indicates that inflammatory processes are closely associated to the pathogenesis of depression (1). Currently, the anti-inflammatory drugs commonly used to treat depression in the clinic are non-steroidal anti-inflammatory drugs (NSAIDs) and Omega-3 fatty acids. NSAIDs, such as aspirin and ibuprofen, are considered to have an antidepressant effect because they can lower inflammation in the body and thus improve depressive symptoms (2). A substantial body of clinical research has demonstrated significant improvements in mood and overall mental health in depressed patients treated with NSAIDs. Omega-3 fatty acids, believed to possess anti-inflammatory properties, have also exhibited favourable outcomes in the management of depression. Studies have indicated that a diet abundant in Omega-3s is associated with a reduced risk of depression. These fatty acids may modulate inflammatory responses and neurotransmitter levels, thereby contributing to their antidepressant effect (3). In addition to the two aforementioned classes of drugs, other substances with anti-inflammatory bioactive effects, such as steroids, have demonstrated antidepressant effects by suppressing the inflammatory response, thereby potentially alleviating depressive symptoms. Patients who have used steroids for a prolonged period have exhibited a substantial reduction in depressive symptoms (4).

Nevertheless, the question of whether these findings can modify the prevailing status quo for depressed patients, who require either long-term or short-term cognitive therapy, remains unresolved (5). This shift in the management of depression emphasises the transition of long-term fixed cognitive therapy into a therapeutic process involving the regular administration of medication. This transformation in the treatment process is undoubtedly more appealing to patient groups requiring short, rapid, and convenient treatments for depression, such as working adults and school students. The present moment is characterised by a crisis in the translation of research findings into clinical practice in the field of depression. This crisis results in the failure to fully implement significant discoveries concerning the underlying mechanisms of depression in clinical treatment. The reasons for this phenomenon are not clear-cut, and there is a pressing need for novel approaches to enhance the return on investment in depression research.

This study proposes that reducing inflammation in the body should be a goal in treating depression. This idea is backed by the new area of research on developing anti-inflammatory drug treatments for depression. Concurrently, the field of artificial intelligence is experiencing rapid growth and gradual infiltration into all aspects of self-health assessment and teletherapy (6,7). In this context, the question arises as to how artificial intelligence can be utilised to assist in the pharmacological treatment of depression under effective human management.

The objective of this scoping review is to examine the current state of inflammation in randomised controlled clinical trial studies of depression. The identification of current trends in this area and the collation of existing frameworks that may be used to guide future research is also intended, with a view to providing new ideas and approaches to the treatment of depression.

## 3 Literature Review

### 3.1 Overview

Neuroinflammation, defined as an immune response within the central nervous system, involves a series of processes including the activation of glial cells and the release of cytokines and chemokines. In the development of depression, abnormal neuroinflammation interferes with neurotransmitter metabolism, affects neuroplasticity, and in turn triggers depressive symptoms. Relatedly, studies using radiomics to predict remission in Cushing’s disease have highlighted the role of inflammation in neuroendocrine contexts, offering insights into similar processes in depression(11).

Conversely, signalling pathways represent a series of intracellular molecular interactions, including the NLRP3 (12), NF-κB (13) and P2X7R (14), which play a pivotal role in the transmission and regulation of neuroinflammation and depression. These signalling pathways are of central interest in the study of the pathogenesis and therapeutic targets of depression. Advanced AI techniques, such as deep learning for segmenting pituitary adenomas, could be adapted to study these signaling pathways in depression (15).

In 2017, Miller and Raison published a image entitled ‘Transmitting stress-induced inflammatory signals to the brain’. This image was edited to reflect the adjustment of factors that trigger inflammation-associated depression from stress to environmental factors, heritage factors, and personal habits **(Figure 1)**. This adjustment is more consistent with existing evidence describing the inflammatory pathogenesis of depression (16). **Figure 1-A** depicts the pathological process by which depression susceptibility factors stimulate inflammation through humoral, neural and cellular pathways. Depression susceptibility factors include environmental factors, genetic factors, and lifestyle habits. The environmental factors referred to here include both human social and family environments and other scenarios where human interaction occurs, as well as natural environments that can trigger microbial infections in humans. Microorganisms are mentioned because patients with depression often have positive lipolyaccharide (LPS) results or signs of infection in their laboratory tests. The strong action of depression susceptibility factors activates the sympathetic nervous system to release catecholamines, and this results in the production of myeloid cells by the bone marrow and their gradual release into the peripheral circulation. Here we describe the inflammatory response to depression initiated by monocytes in a cellular pathway using monocytes in myeloid cells as an example. The monocyte in the peripheral blood circulation will try to fulfil its mission of capture, phagocytosis. Examples include bacterial metabolites or other substances that stimulate damage-associated molecular patterns (DMAPs). The most frequent site in the human body where monocytes are confronted with bacteria and associated metabolites is the gut, where monocytes take full advantage of recognising microbial-associated molecular patterns (MAMPs) and thus clearing microbes. However, DMAPs and MAMPs are optimal mediators for the activation of inflammatory signalling pathways, such that a range of expression occurs within monocytes, such as nuclear factor-κ b (NF-κ b) and pyrin structural domain protein 3 (NLRP3) inflammatory vesicles. This expression is a cascade response that can also interact with each other. For example, stimulation of NLRP3 in turn activates caspase 1, leading to the production of mature interleukin-1β (IL-1β) and IL-18, as well as splitting of the glucocorticoid receptor, leading to glucocorticoid resistance. This expression results in the recruitment of large amounts of cytokines such as IL-1β and IL-18, which in turn recruits other immune cells against the cytokine chemical gradient to come and participate in the immune process. As immune mobilisation continues, the number of cytokines produced gradually increases, and these cytokines do not stop at specific sites of the immune response, but travel with monocytes to the brain via humoral and neural pathways. The immune cells present in the brain are partly called astrocytes and partly called microglia. These microglia are activated by a large number of cytokines to an M1 pro-inflammatory phenotype which releases CC-chemokine ligand 2 (CCL2), which in turn attracts activated monocytes to the brain via the cellular pathway. Once the monocytes enter the brain, the peripheral immune response will spread to the centre, which may result in central inflammation and exacerbate the manifestations of depression. **Figure 1-B** highlights how nerve cells in the inflammatory response affect human behaviour through cellular pathways. If the inflammatory response is present in the brain, then there is no doubt that neurotransmitter transmission and metabolism will certainly be disrupted. The effects we are most interested in are depressive symptoms, such as lack of motivation and lack of pleasure in behaviour. Here elaborate on interferons (IFNs), IL-1β and tumour necrosis factor (TNF) as representatives of cytokines. These three cytokines reduce the availability of monoamine-serotonin (5-HT), dopamine (DA) and norepinephrine (NE) – through activation of the mitogen-activated protein kinase (MAPK) pathway. With cytokine-induced reduction in astrocyte glutamate reuptake and stimulation of astrocyte glutamate release. Glutamate (Glu) is an excitatory amino acid neurotransmitter. Excess Glu, when bound to extrasynaptic N-methyl-D-aspartate receptor (NMDAR), leads to decreased brain-derived neurotrophic factor (BDNF) and excitotoxicity. Low levels of DA and BDNF levels of neurotransmitters passed between nerve cells and neuronal cells are then not consistent with concentration levels in a non-inflammatory state, a result that can lead to a lack of reward motivation and a lack of rapture in cortical striatal circuits.

**Figure 1.**
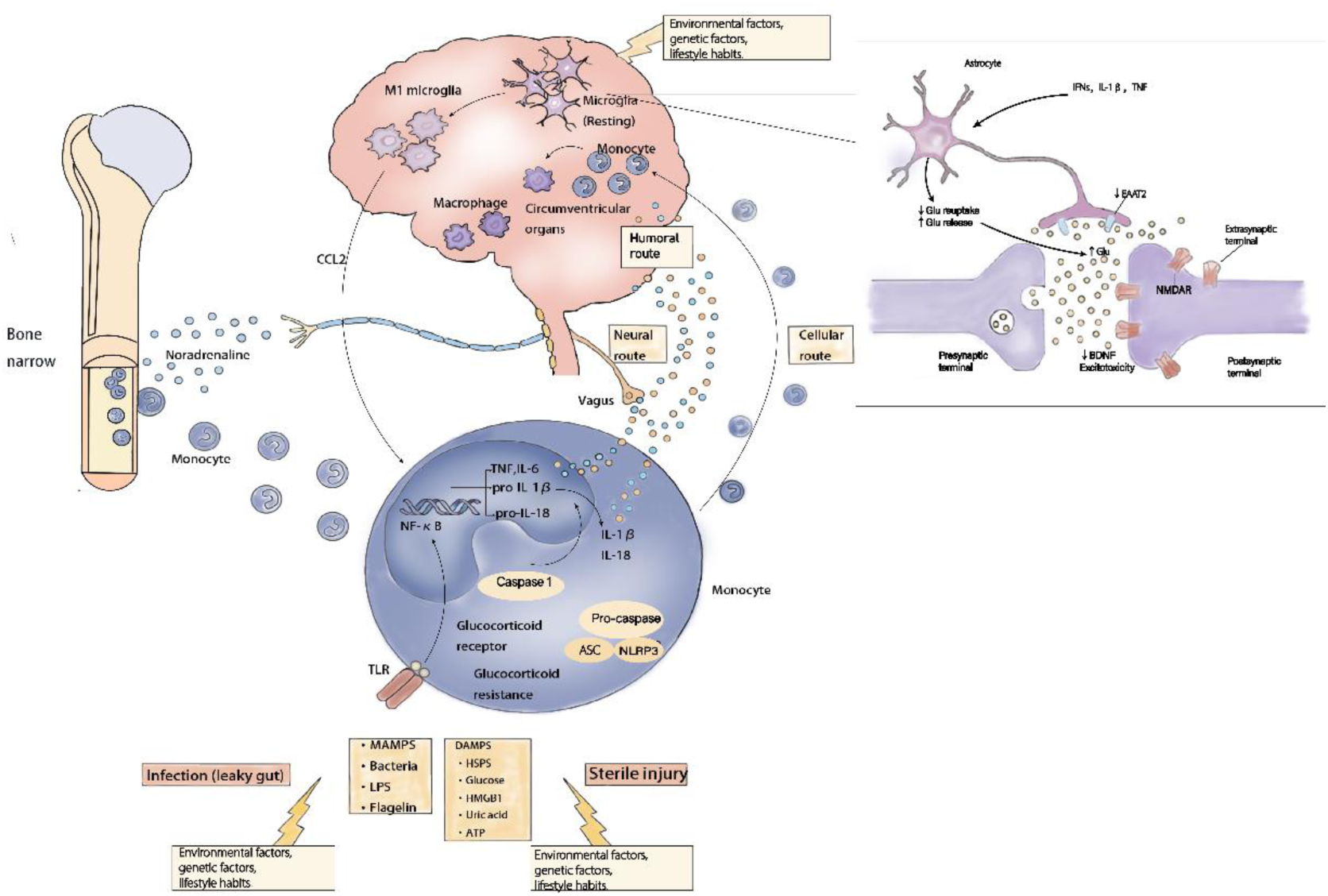
Inflammatory pathogenesis of depression. (A) Humoral, netural and cellular route. (B) Neurotransmitters. ASC, apoptosis-associated speck-like protein containing a CARD; HMGB1, high mobility group box 1; LPS, lipopolysaccharide; TLR, Toll-like receptor. EAAT2, excitatory amino acid transporter 2; NF-κB, nuclear factor-κB; Glu, Glutamate; HSP, heat shock protein; NMDAR, N-methyl-D-aspartate receptor; BDNF, brain-derived neurotrophic factor; Glu, Glutamate; MAPK, mitogen-activated protein kinase; DA, dopamine; NE, norepinephrine; 5-HT monoamine-serotonin; IFNs, interferons; TNF, tumour necrosis factor; CCL2, CC-chemokine ligand 2; IL-1β, interleukin-1β; pyrin structural domain protein 3 (NLRP3); NF-κ b, nuclear factor-κ b; MAMPs, microbial-associated molecular patterns; DMAPs, damage-associated molecular patterns; LPS, lipolyaccharide.

The present review covers 31 animal studies focusing on neuroinflammatory target-based therapeutic strategies for depression, involving both pharmacological interventions (including SGLT2 inhibitors, vitamin D analogues, natural extracts and other compounds) and non-pharmacological interventions (e.g. These include acupuncture, exercise, and light therapy. The present review comprehensively incorporates the results of the animal model studies, with the aim of presenting an all-rounded picture of the research in this field and to provide a comprehensive overview of the clinical translation of the scoping review. It aims to present a comprehensive picture of the research in this field and to provide an animal-level basis for a scoping review to explore clinical translation.

In the actual population of depressed patients, neuroinflammation does not exist in isolation, but rather involves the interaction of multiple systems, including the neurological, endocrine, and immune systems. The lifestyle and environmental factors of each depressed patient have been shown to indirectly affect neural circuits, thereby complicating the manifestation of neuroinflammation. It has been demonstrated that both the patient’s personal factors and the complex neuroinflammation can have a significant impact on the onset, development, and treatment response of depression. The present study has identified a variety of novel anti-inflammatory drugs and non-pharmacological treatments in animal experiments that modulate the three main signalling pathways of neuroinflammation, namely NLRP3 (12), NF-κB (13) and P2X7R (14). This modulation has been shown to result in an improvement of depressive symptoms and neuroinflammation. In terms of pharmacological interventions, Empagliflozin has been shown to regulate autophagy or inflammation and neurogenesis in the hippocampus through activation of AMPK (19). Calcitriol has been demonstrated to inhibit the P2X7R/NLRP3/caspase-1 signalling pathway, thereby reducing neuroinflammation (20). Among non-pharmacological interventions, acupuncture (14) and exercise (21) have been shown to modulate relevant inflammatory and apoptotic pathways, improve neuroplasticity and reduce neuroinflammation, respectively. The primary debate has focused on single-target versus multi-target modulation strategies and pharmacological versus non-pharmacological intervention modalities. Single-target modulation emphasises precise inhibition of specific key signalling pathways, whereas multi-target modulation argues for integrated modulation of multiple associated signalling pathways to cope with disease complexity. The majority of extant studies are in the short-term observation and small-scale research stage. At this stage, animal studies have not yet been able to clearly explain how these drugs act on specific neuroinflammatory targets according to the dynamic changes in animals, and effectively regulate the functional activities of neural circuits in the hippocampus, prefrontal cortex, and other brain regions that are closely related to depression, so as to achieve a stable and long-lasting antidepressant effect.

Although the main focus has centred around therapeutic strategies for depression based on neuroinflammatory targets, the question of how different pharmacological and non-pharmacological treatments can precisely and dynamically modulate the interaction between neuroinflammation and depression-associated neural circuits in a complex human physiological and pathological environment has not been thoroughly investigated.

### 3.2 Development

In the early research on depression, the focus was primarily on the traditional theory of monoamine neurotransmitter imbalance, with a relative paucity of understanding of the role of neuroinflammation in depression (22). However, as research progressed, it was found that the inflammatory response was associated with depression. Nonetheless, at this stage, research remained at the level of observational studies, with no exploration of the specific mechanisms of neuroinflammation-related signalling pathways and effective intervention strategies.

In recent years, however, there has been a shift in focus towards the exploration of the role of neuroinflammation-associated signalling pathways, such as NLRP3 (12), NF-κB (13) and P2X7R (14), in the development of depression. These studies have identified pharmacological and non-pharmacological intervention options with therapeutic potential to modulate these signalling pathways and thereby improve depressive symptoms. However, the majority of these studies are in the animal experimentation phase and have yet to demonstrate sufficient clinical relevance.The findings from these animal experiments are not yet representative of a comprehensive understanding and effective solution to the problem of achieving long-term stable modulation of neuroinflammation-related signalling pathways in depression and avoiding adverse effects in order to achieve a sustained antidepressant effect. This requires a significant volume of in-depth research.

### 3.3 Research status

#### Pharmacological interventions

A study by Muhammad et al demonstrated that Empagliflozin can regulate the balance of autophagy or inflammatory dynamics and PKCζ-mediated neurogenesis in the hippocampus through activation of AMPK, inhibit the NLRP3 inflammatory vesicle pathway, and ameliorate depressive symptoms. In the reserpine-induced rat model of depression, Empagliflozin has been shown to restore monoamines and autophagy or inflammatory balance in the hippocampus, and to promote neuroplasticity. However, no in-depth study has yet been conducted on whether its sustained effects on the signalling pathways are affected by adaptive modulation of the organism after long-term use (19).

A study by Ma et al suggests that the combination of brexpiprazole and fluoxetine produces a rapid antidepressant effect in an inflammatory model of depression, possibly through stimulation of the BDNF-TrkB signalling pathway as well as modulation of dendritic spine density. Consequently, it is hypothesised that brexpiprazole-assisted SSRI treatment is expected to produce rapid antidepressant effects in patients with depression (22).

A study by Bao et al revealed that chronic stress activates endogenous retroviruses (ERVs) in a mouse model, prompting an immune-inflammatory response in microglia in the basolateral amygdala (BLA), which ultimately triggers negative emotional behaviours. Antiretroviral therapy, inhibition of reverse transcriptase, and knockdown of p53, a gene that regulates the transcription of ERVs, have been shown to be effective in suppressing chronic stress-induced immunoinflammatory responses in microglia and ameliorating negative emotional behaviours in mice. This remarkable effect relies on reversing microglial cell morphology and biological immune inflammation. This particular use of antipsychotics may generate new therapeutic inspiration for psychiatrists (23).

Wang et al. demonstrated that vitamin D analogues could attenuate neuroinflammation, alleviate LPS-induced depressive-like behaviours and cognitive deficits, and have protective effects on neurons through inhibition of the P2X7R/NLRP3/caspase-1 signalling pathway. However, the potential adverse effects of long-term use require further investigation. In particular, the indirect effects of abnormal calcium metabolism on neuroinflammation-related signalling pathways need to be further explored (20).

A study by Rostevanov et al. demonstrates that leukotrienes (LTs) enhance depression-like symptoms in mice, exhibiting a non-sex-selective effect. However, this study also reveals that sex selectivity is present in inflammatory homeostasis, as evidenced by the increased levels of IL-6 and TNF-alpha in brain regions in males and decreased levels in females. The clinical and translational implications of this sex-selective correlation in inflammatory homeostasis remain to be elucidated (24).

A study by Li et al. examined the effects of liver-sparing granules (SGKL) on the gut, brain, and behaviour of chronic restraint stress (CRS) rats, and found that SGKL could improve depression. The study also found that SGKL could improve depression-like behaviour by altering the intestinal microbiota and metabolites targeting the PI3K/Akt/mTOR pathway (25).

Progesterone alleviated depressive-like behavior in mice. It did so by reducing neuroinflammatory responses and oxidative stress(26).

A study by Casaril et al. demonstrated that the selenium-containing compound 3-((4-chlorophenyl) selanyl)-1-methyl-1H-indole (CMI) was able to reverse acute restraint stress-induced depressive-like behaviour in mice by regulating oxidative stress and neuroinflammation, and by restoring corticosterone levels and related gene expression. This selenium-enriched compound, endowed with oxidative and anti-inflammatory bioactivities, may represent a promising candidate for future dietary interventions in the prevention of depression (27).

A total of three studies on nutritional supplements have reported that propolis, n-3 polyunsaturated fatty acids (PUFAs) and skeletal muscle glycine all improve depression-like behaviour. This mechanism is associated with anti-inflammatory and antioxidant properties 32−34.

A total of fourteen studies on natural extracts, including Fustin, Berberine, and Cinnamic Acid, were identified as modulating oxidative stress, neuroinflammatory markers, and associated signalling pathways in animal models to enhance depression– and anxiety-like behaviours. However, the majority of these studies were of a short-term nature, and there was an absence of longitudinal research on the stability of the signalling pathways and the potential challenges associated with tolerance during extended use. Furthermore, there is a paucity of long-term follow-up studies.The differences between these studies are summarised in **Table 1**.

**Table 1.**
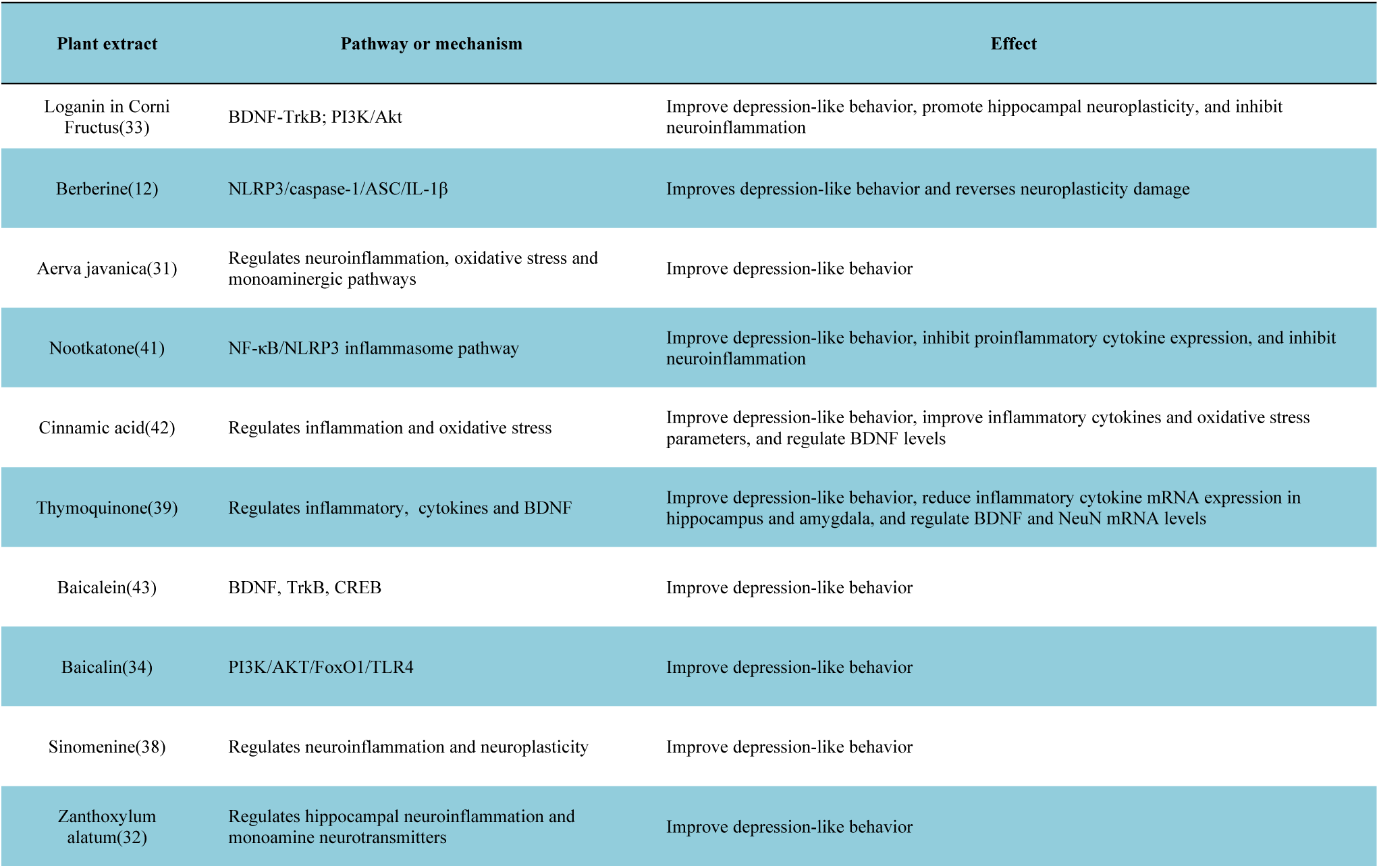
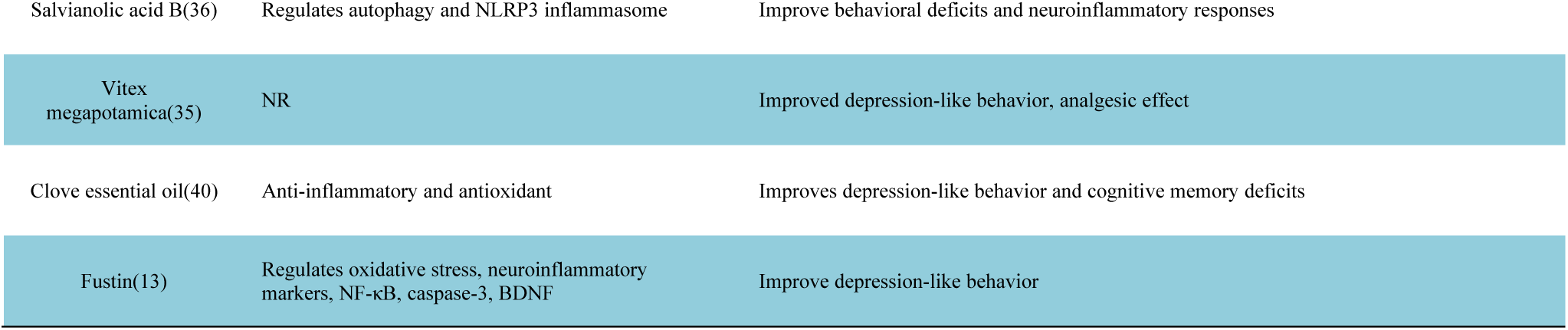
Comparison of 14 research reports on natural extracts. The abbreviations in the table are explained below. BDNF: brain-derived neurotrophic factor; TrkB: tropomyosin receptor kinase B; PI3K: phosphatidylinositol 3-kinase; Akt: Protein kinase B; NLRP3: NLR family pyrin domain containing 3; caspase-1: Cysteinyl aspartate specific proteinase-1; ASC: apoptosis-associated speck-like protein containing a CARD; IL-1β: interleukin-1β; NF-κB: nuclear factor kappa-B; TLR4: toll-like receptor 4; FoxO1: forkhead box O1; CREB: cyclic AMP response element-binding protein; NR: not report.

#### Non-pharmacological interventions

Four studies of acupuncture have indicated that acupuncture modulates oxidative stress, neuroinflammation, and apoptosis-related pathways and attenuates chronic stress-induced depressive-like behaviours and cognitive deficits, but the mechanism of maintenance of the long-term effects of acupuncture stimulation and how individual differences in sensitivity to acupuncture affect the modulation of neuroinflammation-related signalling pathways have not yet been conclusively demonstrated (14,44–46).

A study by Xie et al. from Renmin Hospital of Wuhan University, China, demonstrated that swimming exercise increases the expression of proteins related to neuroplasticity in the hippocampus, reduces neuroinflammation, and improves depressive behaviours. However, further systematic studies are required to investigate the long-term regulation of neuroinflammatory signalling pathways by different exercise intensities, frequencies, and durations, and the differential effects of exercise among individuals of different ages, genders, and health conditions. Further systematic studies are needed to investigate the differential effects of exercise in individuals of different age groups, genders and health conditions (21).

A study by Universitat de Barcelona, Spain, examined the effects of brain-gut photobiomodulation (PBM) on mice with chronic stress. Chronic stress is an important risk factor for depression and is associated with gut microbiota dysbiosis. Existing treatments for depression are deficient, and PBM has potential as a non-invasive therapy. Utilising an experimental model, the study ascertained that chronic stress instigates alterations in behaviour and hippocampal protein expression in mice. Moreover, the study determined that PBM modulates the gut microbiota, restores hippocampal Sirt1 levels, improves cognition and reduces neuroinflammation. This novel physical light therapy approach to treat stress-related depression provides a novel direction for further research (47).

### 3.4 Perspective

The choice of single target versus multi-target regulation is a major point of debate in the field.Some studies have argued in favour of the search for efficient inhibitors targeting specific key signalling pathways. These studies suggest that precise blockade of the pathway can effectively control neuroinflammation and thus improve depressive symptoms. For example, certain natural compounds have been studied with a focus on their inhibitory effects on NLRP3 inflammatory vesicle activation and have achieved some antidepressant effects (12,36,41). Conversely, it has been proposed that depression is a multifactorial and intricate condition, and that single-target modulation may not be sufficiently comprehensive in addressing the multifaceted nature of the disease and the heterogeneity of individual patients. The argument is made that a more effective approach would be the comprehensive modulation of multiple interconnected signalling pathways, such as the concurrent modulation of autophagy, inflammation, and oxidative stress-related pathways, to achieve a more enduring antidepressant effect. Empagliflozin’s ability to simultaneously regulate multiple signalling pathways is considered to be a significant mechanism contributing to its antidepressant effects (19).

A further point of contention for achieving clinical translational value is the comparison of the advantages of pharmacological versus non-pharmacological interventions. Pharmacological interventionists emphasise the precision and actionability of drugs in modulating neuroinflammatory signalling pathways, which can act directly on the target through specific molecular mechanisms and are easy to standardise. Non-pharmacological interventionists counter by highlighting the advantages of natural, holistic regulation and reduced adverse effects that non-pharmacological methods, such as acupuncture and exercise, offer. These methods can indirectly regulate neuroinflammation by improving the physiological and psychological state of the body. From a holistic standpoint, these approaches may be more suited to long-term maintenance therapy. However, the mechanisms underlying non-pharmacological interventions are intricate and challenging to delineate with precision, and the intricacies of their regulation of neuroinflammation-related signalling pathways are yet to be thoroughly investigated.

The present study offers a plethora of novel concepts and methodologies for the management of depression. Nevertheless, considerable progress is yet to be made before the comprehensive resolution of the long-term stable regulation of neuroinflammation-related signalling pathways and the circumvention of adverse effects can be achieved. Future research should focus on the following fuzzy general directions: first, to carry out long-term follow-up studies to observe the long-term effects and potential adverse effects of different interventions in animal models, and to analyse in depth the dynamic changes of the signalling pathways; second, to strengthen the research on individual difference factors (e.g., gene polymorphisms, lifestyle, intestinal flora, etc.) and to explore the personalised treatment options; thirdly, to further explore the different interventions between theThirdly, to further explore the joint application mode and synergistic mechanism between different interventions, to give full play to the advantages of pharmacological and non-pharmacological interventions, and to improve the effect and stability of depression treatment.

It is the contention of this team that the progress made thus far in animal studies is not transferable to a clinical context. With regard to research methodology, there is a necessity for animal models to be further optimised in order to more accurately replicate the pathophysiological processes of human depression. While the development of new animal models for inducing depression is a more arduous task, the enhancement of the design and implementation of clinical studies is a more attainable goal, thereby improving the extrapolation and utility of findings. While acknowledging the value of animal studies in identifying interventions that promote or inhibit key factors, it is crucial to recognise the limitations in assessing the extent to which these interventions occur in actual organisms. It is highly probable that moderation conclusions are minor or insignificant in real-world settings. To enhance the credibility of the research, the authors’ team should consider publishing the specific numbers in addition to the published data and images, rather than relying solely on the P-value significance. The bias caused by the credibility of the interpretation of results by the mere independent presence of P values has been previously reported (48). In addition to the continued exploration of new drug targets and compounds, attention must be paid to the safety and tolerability of drugs, especially with regard to potential risks during long-term use. It is evident that discovering a promising new drug is a very difficult process.

In the context of non-pharmacological interventions, a comprehensive analysis of the mechanisms underlying acupuncture, exercise and other therapeutic modalities is imperative. This analysis should encompass the specific sites and modalities of their regulation of neuroinflammation-related signalling pathways. The objective is to formulate a more scientific and standardised intervention protocol. However, it should be noted that acupuncture is a technique that falls under the umbrella of traditional Chinese medicine (TCM). TCM is an empirical medicine, and the deliberate administration of acupuncture using scientific protocols may potentially diminish the original therapeutic effect. Interdisciplinary cooperation is crucial for comprehensive analysis of the pathogenesis and therapeutic principles of depression, integrating multidisciplinary knowledge such as neuroscience, immunology, psychology, genetics, etc. Unfortunately, in-depth studies from these perspectives can erode research teams’ confidence. The current ability and methods to study from the subcellular organelle level are limited, and compared to the cost of research, cognitive therapy and exhortation to daily exercise may be the most effective option at this time.

## 4 Methods

My previous initial search of the PubMed database revealed no extant systematic reviews publishing insights on how to view anti-inflammatory strategies based on the treatment of patients with depression-related inflammation. Therefore, I conducted a scoping review of extant clinical randomised controlled studies based on anti-inflammatory strategies for the treatment of patients with depression-related inflammation. The purpose of the scoping review was to sort out existing treatment protocols, assess the rationale and consistency of the selection of inflammatory biomarkers, evaluate the value to clinical practice, and make implementable recommendations. This scoping review was previously registered on the OSF platform at (17,18). To ensure sufficient transparency and detail, I followed a standard scoping review framework, first defined by Arksey and O’Malley (49), then later expanded by Levac et al.(50). This review is reported in accordance with the PRISMA extension for scoping reviews (51).

### 4.1 Search strategy and selection criteria

As clinical randomised controlled studies are the highest level of research evidence that constitutes guidance for clinical practice, all strategies involving inflammation-based strategies whether pharmacological or non-pharmacological regimens, but applied to patients with depression-related inflammation, should be included. This is similar to Fei Zhao’s emphasis on the inclusion of randomised controlled studies in evidence-based medical research (5). In this scoping review, depression was recognised by the DSM-V specification(52). Depression-associated inflammation, on the other hand, is not nowadays recognised as a diagnostic term, and therefore inflammation was recognised according to the Mesh entry specification in the Pubmed database (53).

I excluded non-randomised controlled studies, as well as studies that did not diagnose depression purely as depression or co-morbidity. This is because the idea of treating depression based on inflammatory strategies is not yet commonly used in clinical practice and remains in theoretical discussion, and expanding the scope of the disease is not conducive to synthesising the evidence. In addition, I excluded literature published in non-English languages, editorials, conferences, books, advocacy or secondary research that could not be accessed in sufficient detail, which is similar to Lorna’s idea (54).

On 12 November 2024 I performed a systematic search of the PubMed database. This study was designed as a scoping review rather than a systematic review. Therefore, Embase, the Cochrane Library, and Web of Science were not searched due to concerns about the speed and efficiency of the scoping review, despite the fact that multiple databases were searched more broadly and comprehensively.This is similar to the point made by Johannes(55). I did not search Embase, the Cochrane Library, or Web of Science. In my searches, no date constraints were applied in order to be as comprehensive as possible. The complete search strategy is shown below:

1#: (((((“Depression”[Mesh]) OR (Depressive Symptoms[Title/Abstract])) OR (Depressive Symptom[Title/Abstract])) OR (Symptom, Depressive[Title/Abstract])) OR (Emotional Depression[Title/Abstract])) OR (Depression, Emotional[Title/Abstract])

2#: ((((“Inflammation”[Mesh]) OR (Inflammations[Title/Abstract])) OR (Innate Inflammatory Response[Title/Abstract])) OR (Inflammatory Response, Innate[Title/Abstract])) OR (Innate Inflammatory Responses[Title/Abstract])

3#: (((((“Therapeutics”[Mesh]) OR (Therapeutic[Title/Abstract])) OR (Therapy[Title/Abstract])) OR (Therapies[Title/Abstract])) OR (Treatment[Title/Abstract])) OR (Treatments[Title/Abstract])

4#: 1# AND 2# AND 3#

Once the search was completed, I would then import the literature into Zotero Literature Manager for screening. As the scoping review was done independently by me, I screened the literature a total of 3 times in order to avoid screening bias and reviewed the results of each screening, which were in almost perfect agreement. The use of kappa coefficients for measuring the confidence of those who screened the literature is not reported here as it does not involve a second person who screened the literature.

### 4.2 Data charting

Data were extracted for the final included literature and the detailed programme was referred to the research protocol(17).

(1) Basic information such as study authors, year, country, journal / publication source, financial support and type of study.
(2) Patient participation information such as sample size, study subgroups, level of depression, diagnostic criteria, age and gender of test and control groups.
(3) Details of pharmacological or non-pharmacological treatment regimens based on anti-inflammatory strategies.
(4) Study results and lessons learnt.

### 4.3 Data synthesis

This scoping review primarily uses narrative synthesis to superimpose evidence and logical reasoning to create a map of the inflammatory pathology of depression **(Figure 1)**. In case of any implausibility or lack of rigour in the research, I will use logical retrospective reasoning and add appropriate solutions or recommendations.

## 5 Results

### 5.1 Search and study characteristics

Included articles were published between 2008 and 2024 **(Table 2)**. Studies were conducted predominantly in the United Kingdom (n=3) and the China (n=3). Studies pertaining to RCT (n=12) methods were identified **(Figure 2)**.

**Figure 2.**
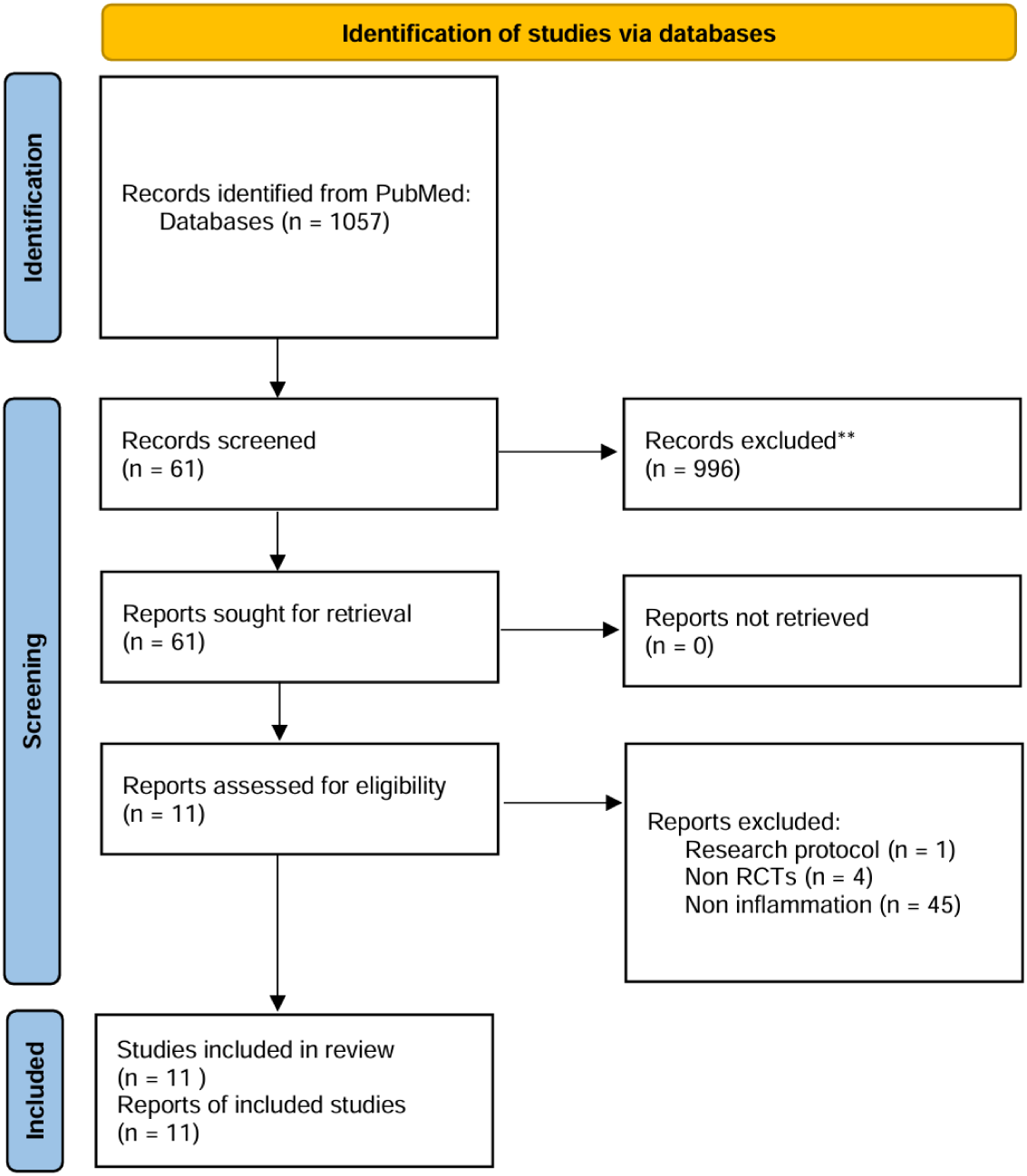
PRISMA flow diagram. A total of 1057 articles were screened. Thirty-two articles met eligibility criteria, including 11 articles. This is represented in the PRISMA flow diagram. Study characteristics were extracted from primary studies only, and all 11 articles were included in the content analysis.

**Table 2.** Study characteristics of included articles (n=11 articles).

| Study author | Year | Country | Journal/source of publication | Funding received | Study type |
| --- | --- | --- | --- | --- | --- |
| Gillian M. Mackay(56) | 2008 | United Kingdom | Clinical and Experimental Pharmacology and Physiology | No | RCT |
| Charles L. Raison(57) | 2012 | United States | JAMA psychiatry | Yes | RCT |
| JeremyF.Weinberger(58) | 2014 | United States | Brain,Behavior,andImmunity | Yes | RCT |
| Zhang Xiaoling(59) | 2018 | China | Pakistan journal of pharmaceutical sciences | No | RCT |
| Chen Mu-Hong(60) | 2018 | Taiwan (China) | Psychiatry Research | Yes | RCT |
| Zhi-Yong Cao(61) | 2020 | China | Journal of Affective Disorders | Yes | RCT |
| Nastaran Hariri(62) | 2020 | Iran | Phytotherapy research: PTR | Yes | RCT |
| Maria Antonietta Nettis(63) | 2021 | United Kingdom | Neuropsychopharmacology | Yes | RCT |
| Bernhard T. Baune(64) | 2021 | Australia | European Neuropsychopharmacology | Yes | RCT |
| Maria Antonietta Nettis(65) | 2023 | United Kingdom | Journal of Psychopharmacology | Yes | RCT |
| Emma Sampson(66) | 2024 | Germany | International Journal of Neuropsychopharmacology | Yes | RCT |

### 5.2 Patient engagement characteristics and identification

The eleven studies included in this review collectively encompassed 907 patients diagnosed with depression, with major depression being the most prevalent form of depression (n=8). The primary diagnostic criterion employed for depression was the DSM-IV (n=7).The majority of the depressed populations studied were middle-aged. The anti-inflammatory drugs employed in the treatment of these patients included Fluoxetine (n=1), Infliximab (n=2), ketamine (n=1), Minocycline (n=2), Votioxetine + celecoxib (n=2), selective serotonin reuptake inhibitors (n=1), Lacquer Tree (n=1). The dispersion in the distribution of these drugs was large **(Table 3)**.

**Table 3.**
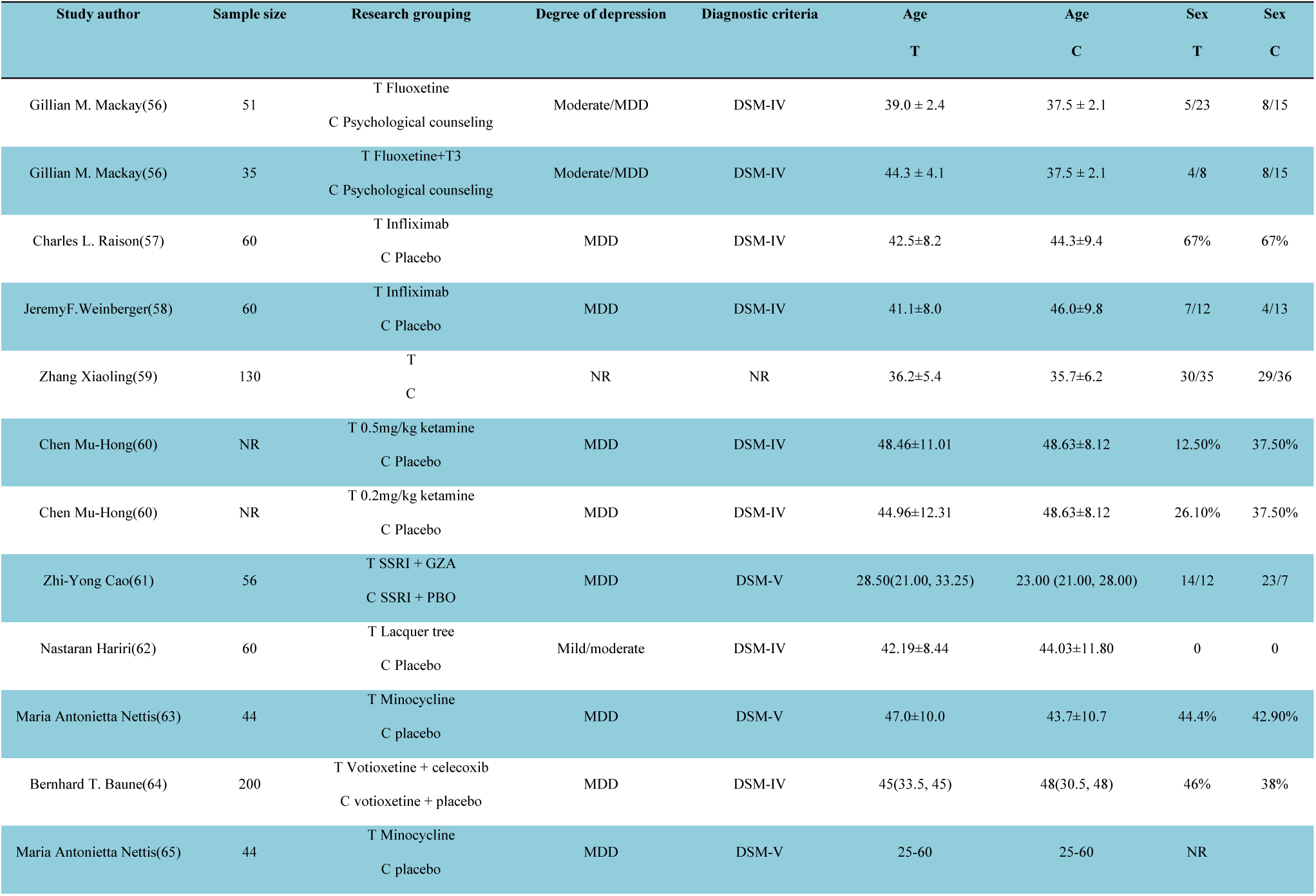
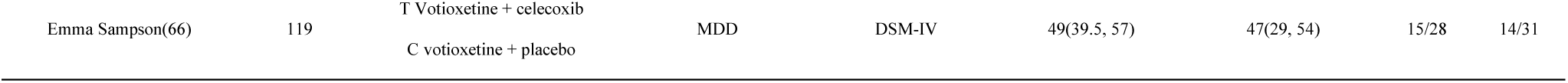
Patient engagement characteristics of included studies (n=11). T indicates experimental treatment method in the study subgroup. C indicates placebo method or other common treatment. MDD indicates major depressive disorder. Sex takes the form of male/female ratios or percentages are shown. *Marked places are for median and interquartile representation. Places marked with # denote minimum and maximum values.NR denotes not reported. DSM-IV and DSM-V denote the fourth and fifth editions, respectively, of the American Psychiatric Association’s Diagnostic and Statistical Manual of Mental Disorders.

### 5.3 Time and inflammatory index

In the eleven studies that incorporated drug treatment cycles ranging from two to eighteen weeks, the vast majority of inflammatory markers examined cumulatively were undetectable in depressed patients. The inflammatory markers that were reasonably detectable in patients with depression in the clinical studies included CRP, hs-CRP, sTNFR1, sTNFR2, IL-2, IL-6, and TNF-α. The utilisation of hs-CRP as an inflammation indicator could be approached via a stratified strategy, e.g., by establishing cut-off levels for hsCRP at 3 mg/L. The benefits of this stratification remain to be elucidated.The merits of such stratified analyses are yet to be demonstrated. A thorough review of the extant literature reveals that the concept of inflammation remains ambiguous in these studies, with the methodology for assessing the dynamic recovery of depression still relying on traditional depression rating scales. Furthermore, there is a paucity of investigation into the concept of inflammation and the indicators of inflammation mentioned in the study’s methodology **(Table 4)**.

**Table 4.**
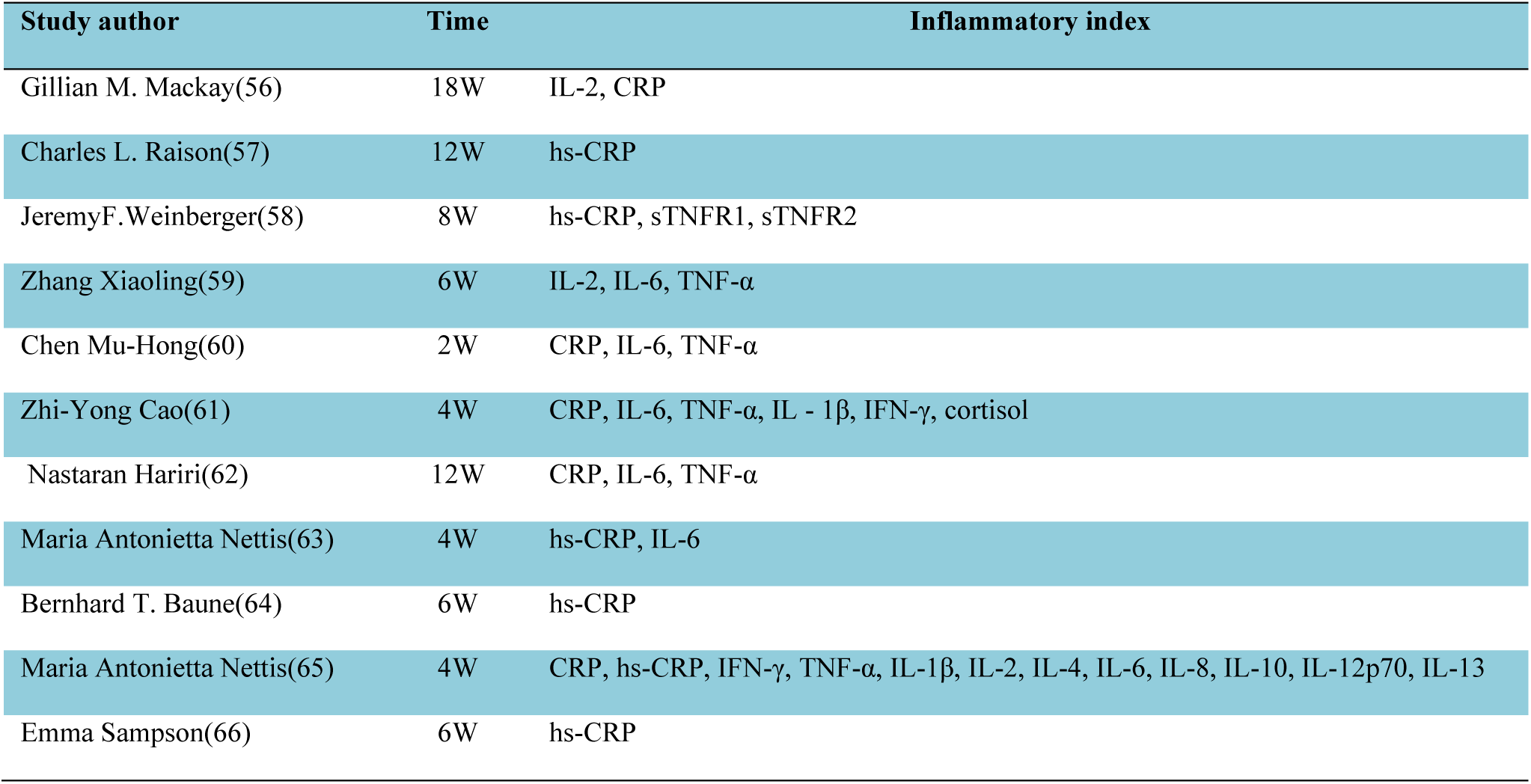
Time and inflammatory index. The following section provides an explanation of the abbreviated terms. IL-2: Interleukin-2; CRP: C-reactive protein; hs-CRP: High-sensitivity C-reactive protein; sTNFR1: Soluble Tumor Necrosis Factor Receptor 1; sTNFR2: Soluble Tumor Necrosis Factor Receptor 2; IL-6: Interleukin-6; TNF-α: Tumor Necrosis Factor-α; IFN-γ: Interferon-γ; IL-1β: Interleukin-1β; IL-4: Interleukin-4; IL-8: Interleukin-8; IL-10: Interleukin-10; IL-12p70: Interleukin-12p70; IL-13: Interleukin-13.

### 5.4 Benefits and challenges of patient engagement

Anti-inflammatory strategies have been shown to have the potential to improve symptoms in the treatment of depression. Studies using glycopyrrolate (GZA) as an adjunctive treatment found that patients treated with an SSRI + GZA had more pronounced remission of depressive symptoms and higher rates of treatment response and remission at 2 and 4 weeks of treatment compared to the placebo group. This finding suggests that by reducing inflammation, it may enhance the mood symptoms of depressed patients and improve treatment outcomes. Anti-inflammatory treatments help to regulate the inflammatory state in depressed patients. A study of overweight or obese depressed women found that supplementation with lacuna (Rhus coriaria L.) in combination with a calorie-restricted diet significantly reduced the patients’ body weight, BMI, body fat, visceral fat levels, and levels of MDA, an indicator of oxidative stress. Additionally, serum levels of IL-6 and TNF-alpha (although not to a significant level) and hs-CRP levels were significantly reduced. This finding suggests that anti-inflammatory strategies may have a positive effect on the inflammatory state of the body in patients with depression by regulating the levels of inflammatory factors, thereby improving the condition.

A significant challenge for precision treatment is the existence of substantial inter-patient variability in response to anti-inflammatory treatments, as evidenced by a study of minocycline in patients with treatment-resistant depression. While no substantial difference was observed in terms of improvement in depressive symptoms between the minocycline and placebo groups, further stratified analyses revealed that patients with baseline hsCRP levels ≥3 mg/L exhibited a more favourable response to minocycline, while those with <3 mg/L demonstrated a less favourable response. This finding underscores the notion that a patient’s baseline inflammation level exerts a significant influence on their response to anti-inflammatory therapy. Consequently, there is an imperative for the development of more precise biomarkers to facilitate the screening of patients who may benefit from personalised treatment regimens. A further critical concern pertains to the safety of anti-inflammatory drugs, which is a pivotal consideration in clinical practice. Some studies have identified potential risks associated with certain drugs. For instance, celecoxib, when studied as an adjunct to the antidepressant vortioxetine, was found to be non-inferior to a placebo in the treatment of depression. However, the drug itself carries a risk of side effects. Therefore, when selecting anti-inflammatory drugs for depression treatment, it is crucial to carefully weigh efficacy against safety to ensure that the benefits outweigh the risks for patients during treatment.

Inflammatory marker levels might be able to predict how a patient will respond to anti-inflammatory treatment. For example, the levels of certain markers like CRP and TNF at the start of treatment have been linked to how well a drug called infliximab works in treating severe depression. Patients with higher starting levels of these markers tend to do better with infliximab, while those with lower CRP levels do better with a placebo. This finding underscores the potential of inflammatory markers as significant predictors of treatment response, thereby aiding in the informed decision-making process for clinical practitioners.

I still don’t fully understand how inflammatory markers affect the treatment of depression, and more research is needed to figure out what they mean and how important they are. One study looked at fluoxetine and escitalopram for treating depression. It found that levels of inflammatory factors like IL – 2, IL – 6, and TNF – α went down a lot in both groups after treatment. But the drop was bigger in the escitalopram group, and this was related to how much the depressive symptoms improved. This finding suggests that distinct inflammatory factors may contribute differently to the pathophysiology of depression and respond variably to treatment.

Further research is necessary to elucidate their mechanisms of action. Anti-inflammatory strategies for treating depression have shown benefits only in cases where patients exhibit high inflammatory factors. However, these strategies also face challenges, such as individual differences and drug safety. Inflammation-related biomarkers have been found to be strongly associated with treatment efficacy. As outlined in **Table 3**, the cost of testing for these inflammatory indicators in a standard hospital in China ranges from 1000 to 1500 RMB, while traditional therapies, based on the experience of our group’s common discussion that 1000 RMB is sufficient to complete a course of depression treatment in any hospital in China. It is evident that the drug regimens employed in conjunction with anti-inflammatory strategies or those intended to calm inflammation are less generalisable.

Future research should address the following questions:

(1) How can we find out if a depressed patient has a strong inflammatory response?
(2) Should the goal of treating depression with anti-inflammatory methods be to stop the inflammation or just to make the depression better?
(3) If a patient takes both regular drugs and anti-inflammatory drugs, how do we classify these drugs?
(4) Do psychiatrists have a clear list of which anti-inflammatory drug combinations to use?
(5) Can using anti-inflammatory drugs help depressed patients take their medicine more regularly and get better faster?

The above five questions should be thoroughly investigated in the development of anti-inflammatory-based treatments for depression, and we call on psychiatrists, neurological surgeons, and psychotherapists to be open and transparent in their research and to publish their findings.

## 6 Discussion

A scoping review was conducted to identify examples of medications based on anti-inflammatory strategies for the treatment of depression. This review yielded a substantial amount of information for future research in this field. While the prevalence of anti-inflammatory strategy-based medications for depression is currently low, the included studies accommodated 907 patients in total, with treatment based on anti-inflammatory strategies exhibiting significant variation.

The findings of the present literature review section revealed that the immune mechanisms of inflammation do not apply to depression. In cellular and animal studies, experimental models and intervention protocols are fixed, which facilitates the placement of confounding factors within a manageable range. The role of immune inflammation in depression is convincing without taking into account the multitude of factors that contribute to the pathogenesis of depression, and is limited to the specific way in which the experimental model is modelled. A particular experimental model of depression itself stimulates depression through inflammatory immune mechanisms (67,68). However, the conditions under which humans acquire depression are uncertain and could be inheritance, environmental changes, acute event stress, etc. (69,70). We hypothesise that some specific triggers of depression are consistent with inflammatory immune mechanisms. Yet, triggers of depression are not usually considered in clinical guidelines for the treatment of depression. Clinical experience within our team has revealed that depression is treated with psychotherapy (the specific type of which depends on the patient’s preferences) when available, and in cases where psychotherapy is ineffective or of low benefit, with antidepressant medication. For patients with depression accompanied by underlying diseases, priority is given to treating the underlying diseases, with psychiatry providing assistance in the synergistic treatment of depression. In the context of depression accompanied by underlying diseases, the hypothesis that immunoinflammation alone can explain the pathological mechanisms of depression is not substantiated. The emergence of anti-inflammatory strategies for the treatment of depression appears to be an endeavour to circumvent the conventional treatment pathway for depression by exploring an alternative inflammatory-immune etiological pathway. A comparison of the potential targets mentioned in the literature review section with the treatment protocols in the randomised controlled studies suggests that this subversive translation is inadequate. Secondly, the new approach to treating depression is much more expensive than the traditional methods. This is because the new pathway may involve more costly drugs, additional tests to monitor inflammation levels, or longer treatment periods, which all contribute to the higher overall cost.

As demonstrated in the literature review section of this paper, the feasibility of treating depression based on anti-inflammatory strategies is derived from inflammatory vesicles (as hypothesised). The hypothesis is that stress activates the NLRP3 inflammasome, which prompts caspase-1 to cleave precursor forms of interleukin-1β (pro-IL-1β) and interleukin-18 (pro-IL-18). The resultant process of NLRP3 inflammasome activation produces the mature inflammatory cytokines IL-1β and IL-18 (71). These cytokines then activate microglia and astrocytes, which, in turn, release interleukin-6 (IL-6), tumour necrosis factor-α (TNF-α), and other molecules (72). This process of increased release of inflammatory mediators (73) could explain why inflammatory indicators in clinical studies frequently refer to IL-1β, IL-6 and TNF-α. However, one of the clinical studies reported that the vast majority of the cytokines were below the detected levels (65), which allows two inferences to be drawn.

The initial hypothesis is that inflammatory vesicles are inadequate in explaining the inflammatory pathogenesis of depression. The secondary hypothesis is that the inflammation indicator is not reasonably set. In order to comprehensively discuss the potential clinical application of the inflammatory pathological mechanism of depression, it is hypothesized that the inflammation indicator is not set up rationally. A salient point is that, according to the activation process of inflammatory vesicles, the reduction of these vesicles in the blood of depressed patients to normal range levels by anti-inflammatory treatment is indicative of the therapeutic endpoint being reached (this set of assumptions is not mentioned in the 11 studies).

It is recommended, based on the assumptions outlined above, that intravenous nano-micro AI robots be used for patients diagnosed with depression, thus addressing the current research deficit in the field. This recommendation is made on the condition that patients are fully informed of the advantages and disadvantages of this intervention, and that the treatment protocol for intravenous nano-mini-AI robots has been approved by an ethics committee following a thorough review. The present state of AI development is constrained by the utilisation of machine learning algorithms that employ vast informational databases stored in remote servers. This technological framework is inadequate for the comprehension of the intricacies inherent in the human body.Nevertheless, the prevailing strategy for addressing depression, which is predicated on anti-inflammatory methodologies, exhibits a dearth of research endeavours concerning the analysis of diverse inflammatory markers within the human organism and the extrapolation of depression treatment outcomes. The integration of an AI robot, capable of being implanted into the human body via intravenous infusion, signifies a potential paradigm shift, promising a future replete with possibilities.

It is imperative to elucidate the following stages of AI development:(1) The production of all inflammatory mediators or cytokines during the activation of inflammatory vesicles is detected using transcriptomics or single-cell sequencing. All potential inflammatory markers detected are then analysed for differences using a healthy adult population and a depressed adult population. This process results in the identification of the first potentially usable inflammatory markers. (2) Considering the rapidity of clinical application, it is necessary to analyse the first batch of inflammation indicators in the blood level again to obtain the second batch of inflammation indicators. (3) Considering the difficulty for clinicians to interpret the data of the inflammation indicators, it is necessary to establish the second batch of inflammation indicators in the healthy adult population to establish a constant model, so that we can get the minimum and maximum values of each inflammation indicator in the healthy adult population. (4) Inflammation indicators that establish a norm in the healthy adult population can be used as the final inflammation indicators. (5) At this stage, there is no accurate and objective assessment method for the diagnosis and evaluation of depression in the medical field, and we need to rely on the subjective judgement of psychiatrists and psychologists. The question of whether the final inflammation index can become the gold standard for assessing depression needs to be addressed by psychiatrists and psychologists rather than psychological rating scales (the protocol for assessing the dynamic treatment effect of depression mentioned in 11 studies is the change in the score of psychological rating scales).

In the context of contemporary clinical randomised controlled studies, the methodology employed for the selection of inflammatory indicators remains largely opaque, thereby engendering a sense of distrust among researchers and practitioners who engage with the extant literature. This opacity hinders the accumulation and advancement of knowledge in the field of clinical translational research on the pathological mechanisms of inflammation in depression. To address these concerns, we propose a systematic approach to the organisation of evidence, commencing from the initial phase delineated above. This approach involves the integration of artificial intelligence (AI) technologies to enhance the transparency and reliability of the research process.

It is hypothesised that the AI could assist researchers, psychiatrists and patients with depression to work together to develop new conceptual therapies based on anti-inflammatory strategies. For example, AI has been successfully applied to improve diagnosis and treatment planning in pituitary adenoma cases, suggesting a model for depression therapy development(74,75).

This hypothesis is based on past experience in developing a shared health needs assessment for the AI integration platform (6). Patients can take information from different test results they get at the hospital and put it into the AI system for analysis(76). The AI system will then carefully look at all the information and keep it up to date as new reports come in. This way, it creates a record of the patient’s health that is focused on the patient, similar to what a hospital would have(77).

The AI integration platform uses closed-source computing on patient data, drawing from a database of all user data on the master server and Internet data (78). This allows patients to easily access their depression health assessment reports without training. It also provides personalized report interpretations. If the report links inflammation to depression, patients can choose anti-inflammatory medication, strengthening treatment beliefs. During treatment, patients can submit medical records to the platform, which projects medication and health changes. Early warning results are sent to doctors, enhancing treatment effectiveness. AI technology improves patient care, reduces medical waste, and helps doctors quickly identify and treat abnormal health data, enabling more precise decisions and better treatment plans.

A research team at Harbin Institute of Technology has developed a micro-robot(79). It can mimic white blood cells, moving through cell gaps to reach blood vessel lumens. The robot can carry medications for targeted treatment, thanks to its internal structure that enables signal transmission for real-time feedback on its internal environment. This real – time feedback is similar to that from imaging and blood tests. The micro – robot has the potential to reduce the economic burden of monitoring inflammation in depressed patients. As the technology advances, it may interact with AI in the future**(Figure 3)**. Wang et al.’s research also supports the proposed AI machine therapy hypothesis(80).

**Figure 3.**
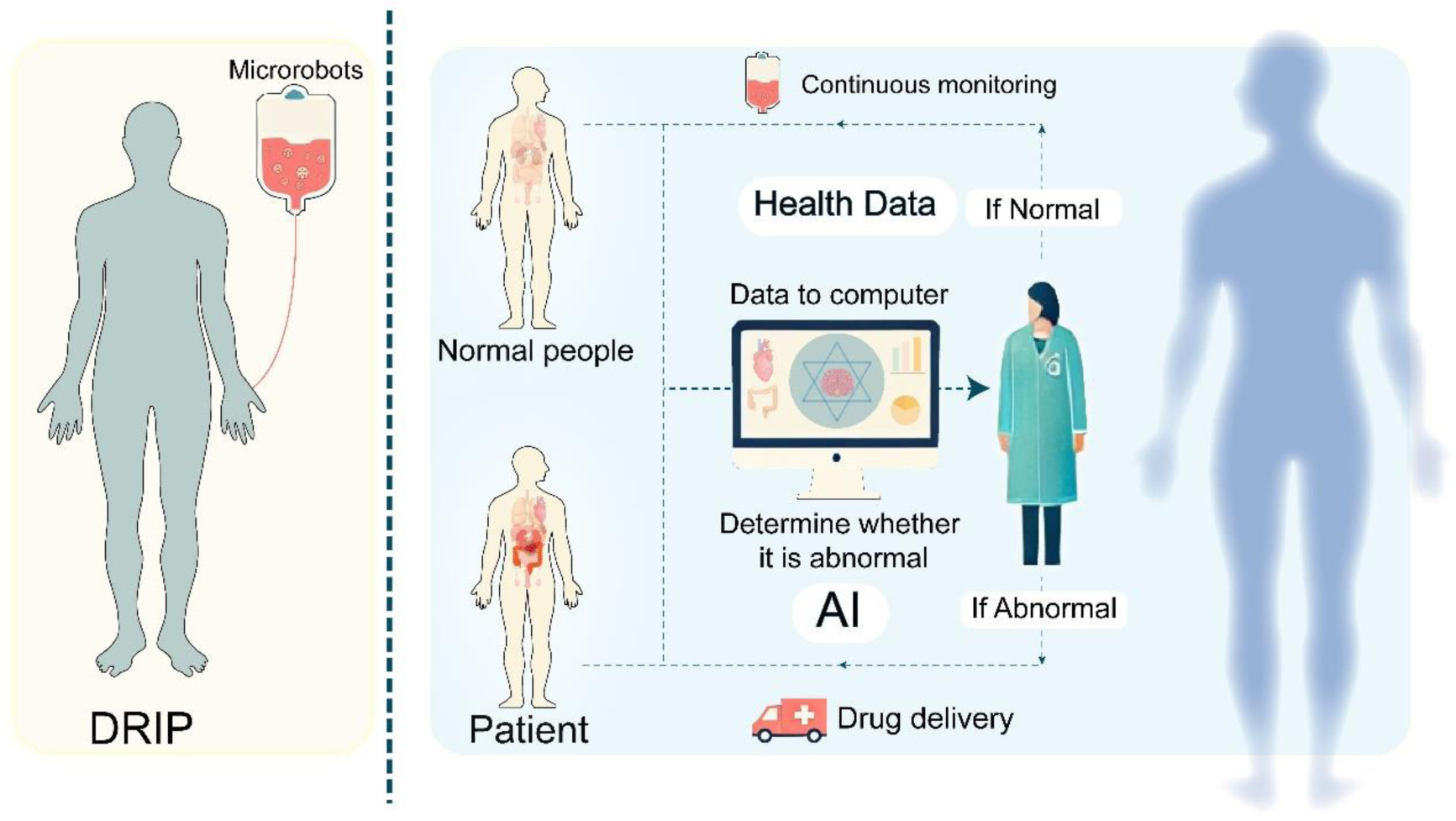
A prospective micro-robot application for interaction with depressed patients. AI microbots treating depression. After patients give consent, microbots are administered into the body. They start a 24-hour continuous body examination, uploading data hourly to the healthcare provider’s computer. When an inflamed area is detected, AI compares it with normal data. If abnormal, microbots dispense medication. If not, they notify the provider and help with treatment until the patient’s data is normal. After recovery, with consent, microbots in suspension re-enter the body. They resume 24-hour, hourly-updated body checks, continuously monitoring the patient.

In addition, existing molecular pathological explanations of depression suggest that abnormalities in the neurochemical metabolism of 5-hydroxytryptamine and dopamine are the primary biological pathogenesis of depression. Such lesions within the brain are usually not easily detectable, even when imaging techniques are used. Here we make use of other brain lesions and difficult to detect diseases (pituitary adenomas). This allows a side-by-side assessment of the need for AI and avoids the bias of depression assessment alone. Zhu et al. reviewed 14 cases of ectopic pituitary adenomas and noted that ectopic pituitary adenomas in the cavernous sinus and suprasellar space were easily misdiagnosed as suspicious intrasellar masses on imaging. Apparently, it is difficult to accurately diagnose ectopic pituitary adenomas even with imaging data. Such a clinical assessment dilemma is a great challenge for clinicians (81). Li et al. reviewed previously treated thyrotropin-secreting pituitary adenomas and proposed the new idea of ‘partial remission of pituitary adenomas alone’.

However, the clinical presentation of pituitary adenoma patients is insidious and difficult to track, especially with the suspicion of a co-secretory tumour (82). How can clinicians input data from pituitary adenoma patients into an AI platform that assesses the disease progression and insidious clinical presentation of patients with pituitary adenomas, and what are the outcomes? It is possible that the poor prognosis of the current pituitary adenoma patient will be known sooner than the human physician. Previously treated suprasellar pituitary adenomas were reviewed by Zhu et al. Nine patients were treated with extended endoscopic transsphenoidal surgery and only one tumour recurred without serious postoperative complications. The implication of this retrospective study is that extended endoscopic transsphenoidal surgery may be a safe and effective method of resecting these tumours. In type II Suprasellar pituitary adenomas, the pituitary stalk should be carefully protected intraoperatively. This protection of the pituitary stalk will be easier to achieve if an AI platform is available to assist the surgeon during the operation (83). Li et al. reviewed 232 previous cases of endocrine pituitary tumour disease in children at Peking Union Medical College Hospital. This would have reduced the presentation of cystic changes or haemorrhage by approximately 25% if the diagnosis had been made early and treated aggressively (84). Based on these four papers, we believe that the current dilemma of how to accurately identify hard-to-detect lesions in the brain cannot be overcome in a short period of time without the aid of AI to assist the judgement of human doctors.

## 7 Conclusion

At present, the use of anti-inflammatory strategies for the pharmacological treatment of depression has limited research value and poor feasibility. The future direction of the new concept of anti-inflammatory strategies for the treatment of depression, proposed in the context of the association between inflammation and depression, is that psychiatrists, researchers, and psychotherapists should shift their future focus from pharmacological treatments based on anti-inflammatory strategies to non-pharmacological treatments of anti-inflammatory strategies, such as positive thinking, exercise, and so on. The popularity of purely clinical randomised controlled studies in the depression population is extremely low when considering the financial investment in research and the benefits of translating the results. In the future, public interest studies, low research costs, and research protocols with mass generalisability will be more likely to stimulate the depression community’s interest in participating in research.

## Supporting information

Scoping review checklist

## Data Availability

All data produced in the present work are contained in the manuscript.

https://osf.io/k9pue/

## 9 Acknowledgments

This scoping review was previously registered on the OSF platform. Pre-established research team members can be accessed in the registration link (https://doi.org/10.17605/OSF.IO/A64GC).

To ensure that authentic authorship qualifications meet authorship standards (https://www.nature.com/nature-portfolio/editorial-policies/authorship). Here, I would like to thank those who have helped in the past in the preliminary stages of this research.

I would like to thank Miss Fengjiao Zhao (Shantou University Medical College), Miss Jingyi Li (North China University of Science and Technology) and Miss Yuanyin Deng (Zhejiang University) for providing assistance in document delivery. And I would like to thank Mr. Yifei Chen (The Affiliated Hospital of Yangzhou University), Hsu Yi Liang (Monash University), Haoran Dai (Nanjing Medical University) and Wenjie Jiang (Northwest Minzu University) for providing assistance in images drawing. Moreover, I would like to thank Miss Ren Sha (Inner Mongolia Chaopai New Materials Co., Ltd.) and Youwei Wang (Beihua University) for providing assistance in management and budget support. What’s more, I would like to thank Mr. Ming Wu (Gongli Hospital affiliated to Second Military Medical University) for providing assistance.

I make sure that the contributions of the collaborators mentioned in this acknowledgement are genuine and valid.

